# Timing of convalescent plasma administration and 28-day mortality for COVID-19 pneumonia

**DOI:** 10.1101/2021.02.02.21250758

**Authors:** Soledad E. González, Lorena Regairaz, Martín R. Salazar, Noelia S. Ferrando, Verónica V. González Martínez, Patricia M. Carrera Ramos, Santiago A. Pesci, Juan M. Vidal, Nicolas Kreplak, Elisa Estenssoro

**Author notes:** **Corresponding autor:** Martin R. Salazar, **Email:**, **Address:** Calle 14 n 320, La Plata, Buenos Aires, Argentina.

## Abstract

Convalescent plasma administration (CPA) is widely used to treat Covid-19, but its effectiveness remains controversial. Here we report the results of an Expanded Access Program of CPA in the province of Buenos Aires, Argentina. We evaluated the relationship between the timing of CPA and 28-day mortality in 4719 hospitalized patients with COVID-19 pneumonia. Early (≤3 days from admission) CPA was associated to decreased mortality in patients in the general ward and in the Intensive Care Unit not requiring mechanical ventilation. This suggests that the favorable effect of CPA might be related both to disease acuity and to the therapeutic window.

## Introduction

Since the beginning of the pandemic by SARS-CoV-2, numerous clinical trials and observational studies have investigated existing therapies which, with the exception of dexamethasone and remdesivir, have proved ineffective ^1,2^. Convalescent plasma has been widely used to treat the disease caused by SARS-CoV-2 (Covid-19) under the hypothesis that it contains potentially neutralizing antibodies against the virus, which can be passively transferred to patients ^3^. The Food and Drug Administration (FDA) has approved the use of convalescent plasma to treat severe COVID-19 patients. In Argentina, in response to the Covid-19 pandemic, the Ministry of Health of the Province of Buenos Aires created the Centralized Registry of Convalescent Plasma Donors (CROCPD-BA) with the aim of collecting, processing and distributing convalescent plasma and issuing recommendations for its use in patients with COVID-19 ^4^.

Despite the potential benefit of convalescent plasma administration, the results of randomized controlled trials or matched–control studies have been controversial ^5-13^, which might be ascribed to differences in disease severity, comorbidities, concurrent treatments, convalescent plasma antibody titers, and timing of its administration, among the reports. While early administration could block the entry of SARS-CoV-2 into the cell and prevent the progression of the disease, deferred administration could be less effective because of intracellular location of the virus and/or end-organ damage due to cytokine storm.

Consequently, the aim of this study was to evaluate the relationship between the timing of convalescent plasma administration and 28-day mortality, in patients hospitalized for moderate to severe COVID-19.

## Methods

This was a multicenter cohort study of data prospectively collected in the National Vigilance System (SNVS 2.0), the Provincial Hospital Bed Management System, and the CROCPD-BA. It includes consecutive hospitalized patients ≥18 years, diagnosed with SARS CoV-2 with RT-PCR, incorporated into an Expanded Access Program of Convalescent Plasma Administration.

Recorded variables were age, gender, comorbidities (arterial hypertension, diabetes, preexistent cardiovascular disease, chronic obstructive pulmonary disease, immunodeficiency), requirement of mechanical ventilation, treatments, and outcomes, as 28-day mortality or discharge. Severe adverse events related to plasma infusion, as transfusion-related acute lung injury (TRALI) and transfusion-associated circulatory overload (TACO) were also registered.

Further data about plasma collection are shown in the Supplement.

The request of convalescent plasma was made by assistant physicians as part of the program, in patients with COVID-19 pneumonia, which was defined as the presence of lung infiltrates, plus one of the following: dyspnea with respiratory rate ≥ 30 breaths/minute, Oxygen saturation ≤93%, Oxygen requirement, PaO2FIO2 <300 mmHg, increase in lung infiltrates >50% during the previous 24-48 hours, alteration in consciousness, multiple organ dysfunction, age >65 years, or any of the aforementioned comorbidities

All units of transfused convalescent plasma had an Ig-G antibody titer ≥1:400, with a volume per unit of 200-250 ml.

Initial severity of illness was assessed according to the hospital site of admission: general ward (GW), Intensive Care Unit (ICU), and ICU admission with requirement of mechanical ventilation (ICU-VM).

We registered the timing of plasma administration with respect to the moment of hospital admission, as < 3 days, between 3-7 days, and beyond 7 days.

The main outcome variable was 28-day mortality. Deaths due to COVID-19 were confirmed on patient death certificates.

Statistical analysis: Continuous variables were expressed as mean ± standard deviation (SD) or median, [0.25-0.75] percentiles. Categorical variables were expressed as percentages. Differences between survivors and nonsurvivors, and between severity on admission and timing of plasma administration were analyzed with chi-square, t, or Mann-Whitney U-tests, as appropriate.

To identify independent predictors of 28-day mortality, variables differing between survivors and non-survivors with a p value <0.10 were entered into a multivariable regression model, using a forward stepwise analysis. For the timing of administration, infusion after 7 days was considered the reference category. Unadjusted and adjusted risks were expressed as odd ratios (OR) and 95%confidence intervals [CI95%].

A two-tailed p value <0.05 was considered significant. Data were analyzed with SSPS-21 (Amonk, NY, US). This study was approved by the Central Ethics Committee of the Ministry of Health of Buenos Aires Province (2020-14965594). The administration of convalescent plasma required signed consent from each patient or legal representative, according to CROCPD-BA regulations (2919/2123/2020).

## Results

In the present study, we analyze 4719 patients with COVID-19 pneumonia admitted to 215 hospitals and treated with convalescent plasma. Epidemiological data of the cohort and comparisons between survivors and non-survivors are shown in Table S1. In univariable analysis, older age, hypertension, diabetes, history of cardiovascular disease (CVD) and chronic obstructive pulmonary disease (COPD) were associated with higher 28-day mortality.

Convalescent plasma was administered to 3036 (64.3%) patients in GW, to 1171 (24.8%) patients in the ICU, and to 512 (10.8%) patients in the ICU-MV subgroup (Table 1). Twenty-eight-day mortality was 22.7% for the entire group; 14.3%, 31.2% and 50.6% for GW, ICU, and ICU-MV patients, respectively (p <0.001). Convalescent plasma was administered to 3113 (66%) patients within the first 3 days of hospital admission, to 1380 (29.2%) patients between days 3-7, and to 226 patients after 7 days; 28-day mortality was 18.1%, 30.4% and 38.9%, respectively (p <0.001).

**Table 1.**
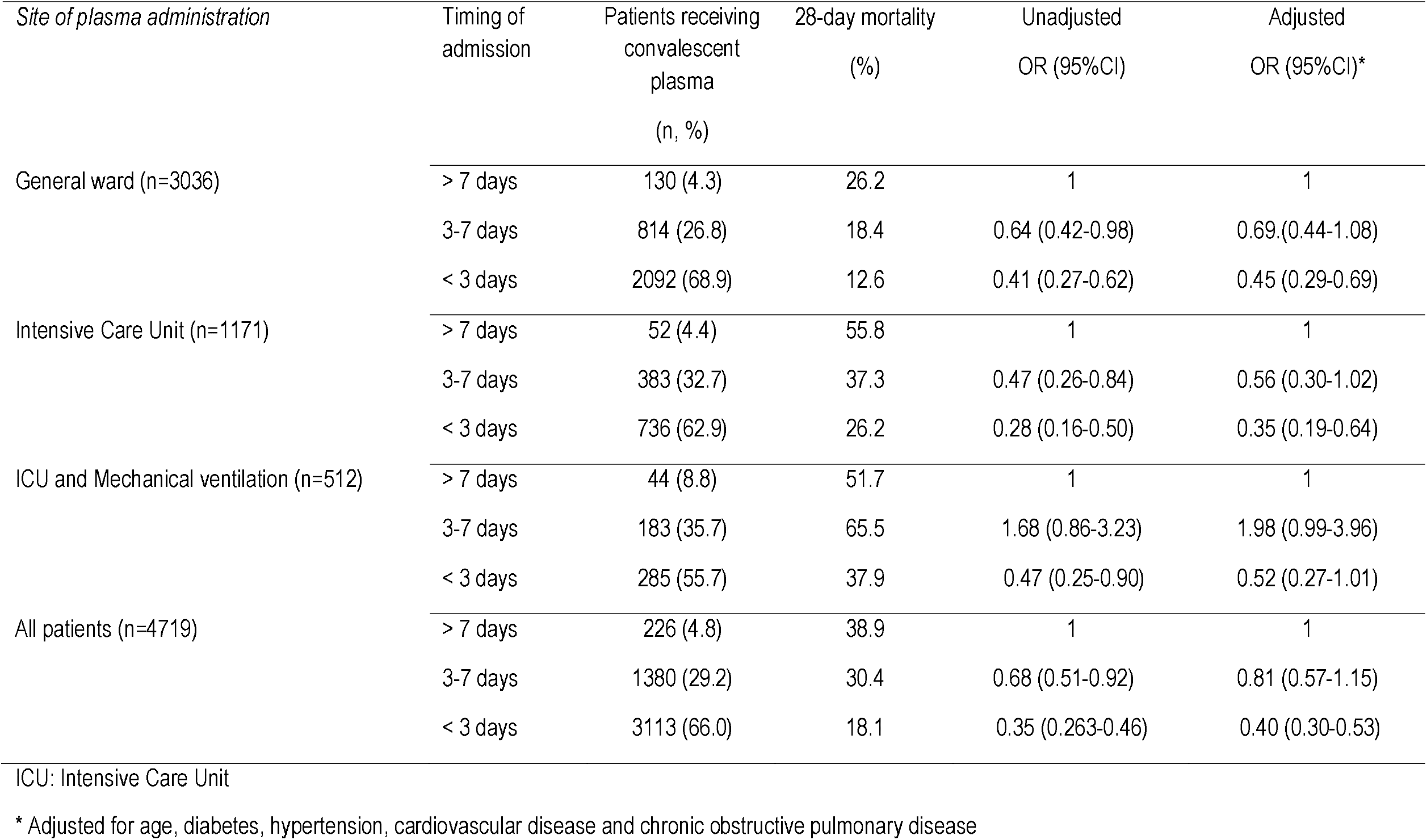
Effect of convalescent plasma on mortality according site of admission and timing of plasma administration.

| *Site of plasma administration* | Timing of admission | Patients receiving convalescent plasma<br>(n, %) | 28-day mortality (%) | Unadjusted OR (95%CI) | Adjusted OR (95%CI)* |
| --- | --- | --- | --- | --- | --- |
| General ward (n=3036) | > 7 days | 130 (4.3) | 26.2 | 1 | 1 |
|  | 3-7 days | 814 (26.8) | 18.4 | 0.64 (0.42-0.98) | 0.69 (0.44-1.08) |
|  | < 3 days | 2092 (68.9) | 12.6 | 0.41 (0.27-0.62) | 0.45 (0.29-0.69) |
| Intensive Care Unit (n=1171) | > 7 days | 52 (4.4) | 55.8 | 1 | 1 |
|  | 3-7 days | 383 (32.7) | 37.3 | 0.47 (0.26-0.84) | 0.56 (0.30-1.02) |
|  | < 3 days | 736 (62.9) | 26.2 | 0.28 (0.16-0.50) | 0.35 (0.19-0.64) |
| ICU and Mechanical ventilation (n=512) | > 7 days | 44 (8.8) | 51.7 | 1 | 1 |
|  | 3-7 days | 183 (35.7) | 65.5 | 1.68 (0.86-3.23) | 1.98 (0.99-3.96) |
|  | < 3 days | 285 (55.7) | 37.9 | 0.47 (0.25-0.90) | 0.52 (0.27-1.01) |
| All patients (n=4719) | > 7 days | 226 (4.8) | 38.9 | 1 | 1 |
|  | 3-7 days | 1380 (29.2) | 30.4 | 0.68 (0.51-0.92) | 0.81 (0.57-1.15) |
|  | < 3 days | 3113 (66.0) | 18.1 | 0.35 (0.263-0.46) | 0.40 (0.30-0.53) |

ICU: Intensive Care Unit
\* Adjusted for age, diabetes, hypertension, cardiovascular disease and chronic obstructive pulmonary disease

The administration of convalescent plasma within the first 3 days of admission reduced 28-day mortality by 60%, compared with the administration after 7 days, and adjusted for possible confounders (Table 1). Absolute values of 28-day mortality according to the timing of plasma infusion in the different strata of severity, reflected by admission site, are shown in Figure 1.

**Figure 1 legend:**
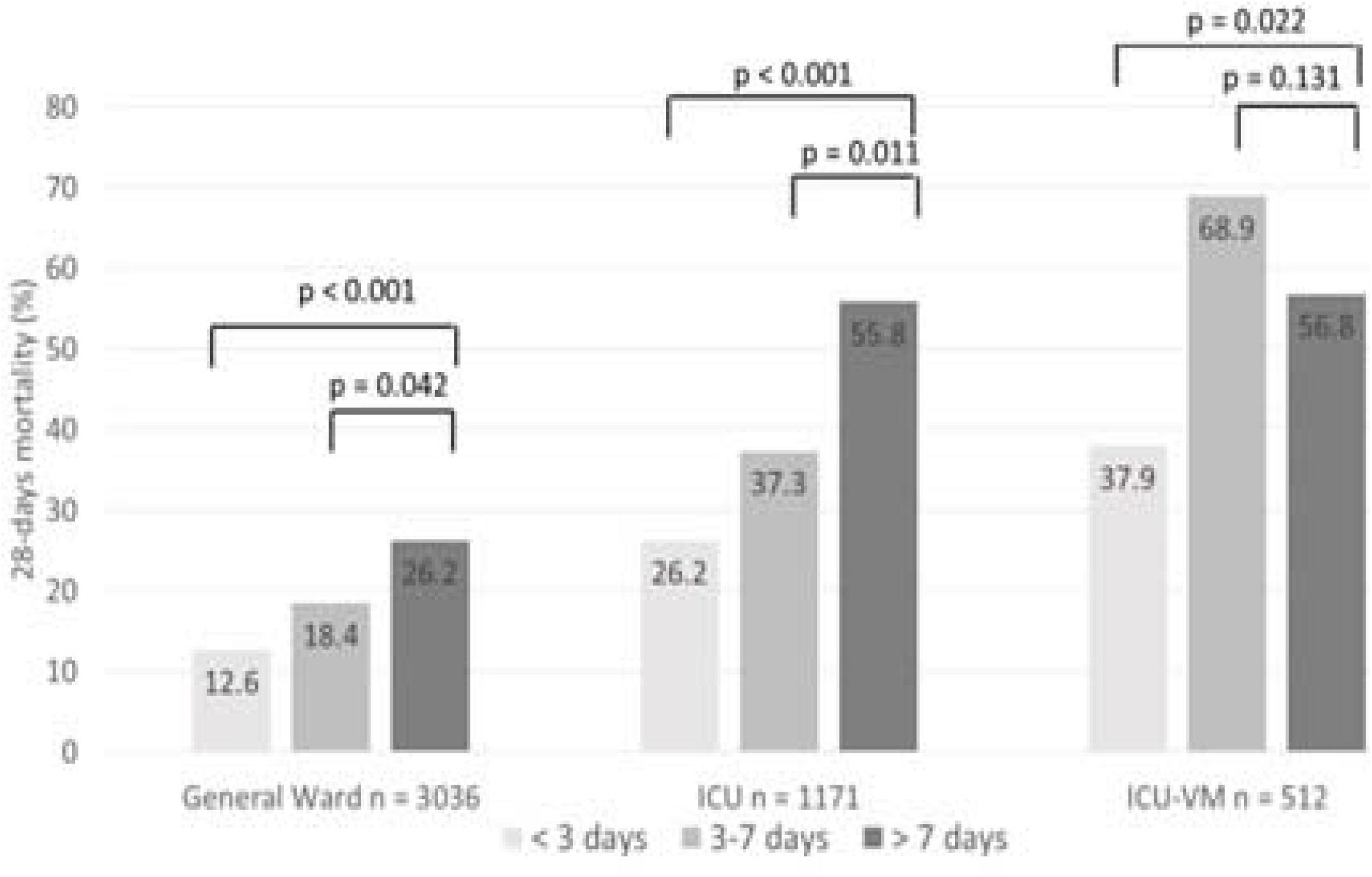
Absolute values of 28-day mortality according to the timing of plasma infusion in the different strata of severity, general ward (GW), Intensive Care Unit (ICU), and ICU admission with requirement of mechanical ventilation (ICU-VM)

The unadjusted and adjusted ORs for 28-day mortality for the different timing groups of convalescent plasma administration, and also analyzed according to admission sites, are shown in Table 1. Adjusted for possible confounders as age, diabetes and CVD, the administration of convalescent plasma within first 3 days of hospital admission was associated with lower mortality in GW and ICU patients not requiring mechanical ventilation. Data regarding antibody titers in the transfused plasma units were available for 2451 patients; 915 (37.3%) had titers ≥1/1600. There were no differences in 28-day mortality in patients treated with plasma units with titers ≥ 1/1600 vs < 1/1600 (23.8% vs. 22.4%, respectively; = 0.416). No episodes of TRALI or TACO were registered.

## Discussion

The most important finding of this study was that the administration of convalescent plasma within 3 days of hospital admission in patients with COVID-19 admitted to the general ward, or to the ICU without need of mechanical ventilation, was associated to a decrease in 28-day mortality. This effect persisted after adjusting for possible confounders as age and comorbid conditions.

The efficacy of convalescent plasma remains controversial. In the two largest clinical trials published to date, a favorable effect of convalescent plasma on different patient outcomes, compared to those who did not receive it, could not be demonstrated. For example, Li et al. did not find any difference in time to clinical improvement between groups ^5^. Yet in 93.9% of patients receiving the intervention, the median time elapsing between symptom onset and randomization was >14 days. Likewise, in the study of Simonovich et al., convalescent plasma was administered at a median time of 8[5-10] days, from the onset of Covid-19 symptoms to enrollment ^10^. A lack of effect of convalescent plasma due to delayed administration cannot thus be discarded. Libster et al. conducted a clinical trial in patients with mild COVID-19, demonstrating that the administration of convalescent plasma with antibody titers higher than 1:1000 within 72 hours of symptom onset halted the progression to more severe disease ^11^. Our results are in line with this last study, underscoring the relevance of early administration of convalescent plasma to COVID-19, and expanding the favorable effect to hospitalized patients admitted to the ward and to the ICU–not requiring mechanical ventilation. Joyner et al and Salazar et al. also reported decreased mortality in a similar group of patients ^12,13^. The benefit, however, was restricted to the administration of convalescent plasma with high titers of anti–SARS-CoV-2 IgG antibody levels. We did not find any relationship between antibody titer levels and mortality.

This study has limitations, mostly due to its observational nature. Unmeasured confounders such as other risk factors or treatments might have influenced the results. Given that severity of illness on admission could not be evaluated with an established score, misclassification of patients might have occurred. Notwithstanding this, the use of admission site as a surrogate of acuity has been already been utilized ^2^; and recently, the rate of clinical improvement after plasma administration could be determined according to the hospital site where the patients received the infusion, among other variables ^15^. A more detailed analysis of the clinical variables collected could not be done, because of the type of registry. Finally, the date of symptom beginning might be a more adequate variable than the time to receiving plasma since hospital admission.

## Conclusion

Our study suggests that the early (≤3 days from hospital admission) administration of convalescent plasma to patients with COVID-19 pneumonia is critical to obtain therapeutic benefit. A 28-day lower mortality was observed both in patients admitted to the general ward and to the ICU, not requiring mechanical ventilation.

## Data Availability

All data referred to in the manuscript is available

## Authorship

All authors fulfill authorships criteria and approved the final version of this manuscript.

LR, NF, PC performed the research; VG, NK, EE and MS designed the research study; SP, JV and NF contributed essential tools; EE, SG and MS analysed the data; SG, EE and MS wrote the paper.

## References

1 Kim MS, An MH, Kim WJ, Hwang TH. Comparative efficacy and safety of pharmacological interventions for the treatment of COVID-19: A systematic review and network meta-analysis. PLoS Med. 2020. http:doi:10.1371/journal.pmed.1003501

2 RECOVERY Collaborative Group, et al. Dexamethasone in Hospitalized Patients with Covid-19-Preliminary Report. N Engl J Med. 2020. http:doi:10.1056/NEJMoa20214

3 Wooding DJ, Bach H. Treatment of COVID-19 with convalescent plasma: lessons from past coronavirus outbreaks. Clin Microbiol Infect. 2020;26(10):1436–1446. http:doi:10.1016/j.cmi.2020.08.005.

4 Gobierno de la Provincia de Buenos Aires. Emergencia Sanitaria. Registro Único de Donantes de Plasma Convaleciente de la Provincia de Buenos Aires (RUDPCBA) para la obtención, procesamiento, distribución y recomendaciones terapéuticas sobre su uso en el tratamiento de pacientes con COVID-19. https://portal-coronavirus.gba.gob.ar/es/efectores-de-salud. Accessed January 21 2020

5 Li L, Zhang W, Hu Y, Tong X, Zheng S, Yang J, et al. Effect of Convalescent Plasma Therapy on Time to Clinical Improvement in Patients With Severe and Life-threatening COVID-19: A Randomized Clinical Trial. JAMA. 2020;324(5): 460–470. http:doi:10.1001/jama.2020.10044

6 Rasheed AM, Fatak DF, Hashim HA, Maulood MF, Kabah KK, Almusawi YA.et al. The therapeutic potential of convalescent plasma therapy on treating critically-ill COVID-19 patients residing in respiratory care units in hospitals in Baghdad, Iraq. Infez Med. 2020;28(3):357–366.

7 Avendano-Sola C, Ramos-Martinez A, Munez-Rubio EC, Ruiz-Antoran B, Malo de Molina R,Torres F, et al. Convalescent Plasma for COVID-19: A multicenter, randomized clinical trial. medRxiv [Preprint] Aug 26, 2020 [cited 2021 Jan 18]. https://www.medrxiv.org/content/10.1101/2020.08.26.20182444v2. https://doi.org/10.1101/2020.08.26.20182444

8 Agarwal A, Mukherjee A, Kumar G, Chatterjee P, Bhatnagar T, and Malhotra P; PLACID Trial Collaborators. Convalescent plasma in the management of moderate covid-19 in adults in India: open label phase II multicentre randomised controlled trial (PLACID Trial). BMJ. 2020;371:m4232. http:doi:10.1136/bmj.m4232

9 Gharbharan A, Jordans C, Geurtsvankessel C, Hollander J, Karim F, Mollema F, et al. Convalescent plasma for COVID-19. A randomized clinical trial. medRxiv [Preprint] Jul 1, 2020 [cited 2021 Jan 18]. https://www.medrxiv.org/content/10.1101/2020.07.01.20139857v1. https://doi.org/10.1101/2020.07.01.20139857

10 Simonovich V, Burgos Pratx L, Scibona P, Beruto M, Vallone MG, Vázquez C,et al; PlasmAr Study Group. A Randomized Trial of Convalescent Plasma in Covid-19 Severe Pneumonia. N Engl J Med. 2020. http://doi:10.1056/NEJMoa2031304. Epub ahead of print.

11 Libster R, Pérez Marc G, Wappner D, Coviello S, Bianchi A, Polack FP; Fundación INFANT–COVID-19 Group. Early High-Titer Plasma Therapy to Prevent Severe Covid-19 in Older Adults. N Engl J Med. 2021. http://doi:10.1056/NEJMoa2033700. Epub ahead of print.

12 Joyner MJ, Carter RE, Senefeld JW, Klassen SA, Mills JR, Johnson PW, et al Convalescent Plasma Antibody Levels and the Risk of Death from Covid-19. N Engl J Med. 2021. http://doi:10.1056/NEJMoa2031893 Epub ahead of print

13 Salazar E, Christensen PA, Graviss EA, Nguyen DT, Castillo B, Chen J et al. Treatment of Coronavirus Disease 2019 Patients with Convalescent Plasma Reveals^i^ Salazar E, Christensen PA, Graviss EA, Nguyen DT, Castillo B, Chen J et al. Treatment of Coronavirus Disease 2019 Patients with Convalescent Plasma Reveals a Signal of Significantly Decreased Mortality. Am J Pathol. 2020;190(11):2290–2303. http://doi:10.1016/j.ajpath.2020.08.001.

14 Shenoy AG, Hettinger AZ, Fernandez SJ, Blumenthal J, Baez V. Early mortality benefit with COVID-19 convalescent plasma: a matched control study. Br J Haematol. 2021. http://doi:10.1111/bjh.17272. Epub ahead of print.

15 Ma T, Wiggins CC, Kornatowski BM, Hailat RS, Clayburn AC, Guo W et al. [Preprint] Jan 20, 2021 [cited 2021 Jan 25]. https://medRxiv2021.01.19.21249678; https://doi.org/10.1101/2021.01.19.21249678

