## Supplemental Table 1 for "Timing of convalescent plasma administration and 28-day mortality for COVID-19 pneumonia"

**Table S1.** Characteristics of the entire group, and comparison between survivors and non-survivors.

|  | All  n = 4719 | Survivors  n = 3647 | Non-survivors  n = 1072 | P  value | 28-day mortality  (%) | Unadjusted OR (95%CI) |
| --- | --- | --- | --- | --- | --- | --- |
| Age (years) | 58 ± 14 | 56 ± 14 | 64 ± 12 | < 0.001 |  |  |
| Gender (male) | 3024 (64.1) | 2317 (63.5) | 707 (66.0) | 0.147 | 23.4 | 1.11 (0.96-1.28) |
| Hypertension | 2017 (42.7) | 1460 (40.0) | 557 (52.0) | < 0.001 | 27.6 | 1.62 (1.41-1.86) |
| Diabetes | 1306 (27.7) | 954 (26.2) | 352 (32.8) | < 0.001 | 27.0 | 1.38 (1.19-1.60) |
| Obesity | 1900 (40.3) | 1457 (40.0) | 443 (41.3) | 0.420 | 23.3 | 1.06 (0.92-1.22) |
| Cardiovascular disease | 546 (11.6) | 364 (11.0) | 183 (17.0) | < 0.001 | 33.3 | 1.84 (1.52-2.24) |
| Chronic obstructive pulmonary disease | 437 (9.3) | 300 (8.2) | 137 (12.9) | < 0.001 | 31.4 | 1.64 (1.32-2.03) |
| Immunodeficiency | 112 (2.4) | 83 (2.4) | 25 (2.9) | 0.417 | 25.9 | 1.19 (0.78-1.83) |

Variables are expressed as mean ± standard deviation, or n (%)

.
