## Supplemental Data for "Timing of convalescent plasma administration and 28-day mortality for COVID-19 pneumonia"

**Supplemental Information**

Early in the COVID-19 pandemic, the Ministry of Health of the Province of Buenos Aires, Argentina created the Centralized Registry of Convalescent Plasma Donors (*CROCPD-BA*).

Possible donors were contacted from patients recovered from COVID-19, on the basis of solidarity and social responsibility principles. After they expressed the will to donate convalescent plasma, donors were scheduled to the centers authorized by the CROCPD-BA, where they signed a written informed consent.

A negative RT-PCR test for SARS-CoV-2 was a prerequisite for donating plasma.

Plasma was obtained by the apheresis method.

Levels of IgG anti-SARS-Cov2 were tested in all units by means of the test ELISA COVIDAR IgG, (Instituto Leloir, Argentina). This test utilizes the trimer of native protein S and a receptor binding domain as antigens, obtained by recombinant DNA techniques produced in human cells.

Antibody titrating was performed in the Immunoserology Section of Central Laboratory of the Children’s Hospital in La Plata, Buenos Aires, Argentina. All units of transfused convalescent plasma had an Ig-G antibody titer ≥1:400.

During the study period (5/14/20 to 10/31/20) 6082 units were transfused (1,09 units per patient on average). The infused volume per unit was 200-250 ml. Dosing was estimated according to weight; patients with <70kg received 1 unit, and those >70 kg received 2 units.
